# Sacral nerve stimulation is more efficient in facilitating pelvic floor responses than pudendal nerve stimulation

**DOI:** 10.64898/2026.10.26.26366499

**Authors:** Mark S. Nissen, Mackenzie B. Moore, Po-Ju Chen, Brinton Vandegrift, Priyanka Gupta, Tim M. Bruns

## Abstract

**Introduction:** Sacral neuromodulation (SNM) is a third line treatment for pelvic organ dysfunction. Pudendal nerve stimulation (PNS) uses SNM electrode leads in an off-label fashion to also treat pelvic dysfunction. Differences in how stimulation of these two nerve targets activates the urethra and external anal sphincter (EAS) may provide insights into the mechanisms of these therapies.

**Materials and Methods:** During SNM lead implant surgeries, needle electrodes and a high-resolution manometry catheter were used to record EAS electromyography (EMG) and multi-segment urethra pressure, respectively. Intra-operative sacral nerve stimulation (SNS) was performed, and EMG and pressure data were analyzed to examine relationships between the responses and stimulation.

Pressure data was then compared to previously collected PNS-evoked responses.

**Results:** Nine participants had data recorded during a SNM lead implant surgery. The applied current threshold to activate the urethra was significantly higher than visual motor threshold (VMT; p = 0.04). The EAS threshold was higher but not significantly different from VMT (p = 0.06). Maximum urethra pressure changes during SNS were not different from PNS driven changes (p = 0.09). Current amplitudes required to elicit maximum changes in urethra pressure were lower for SNS than PNS (p = 0.01) and the maximum change in urethra pressure per mA of applied stimulation was higher for SNS than PNS (p = 0.005).

**Conclusion:** This is the first work to compare muscle recruitment between SNS and PNS in humans. SNS has greater pelvic floor recruitment per step in current than PNS. Typical SNM leads are closer to the nerve target, which may contribute to this effect.

## 1 Introduction

Minimally invasive surgical techniques are often used as a treatment following failure of oral medication for overactive bladder (OAB) and pelvic floor dysfunctions (PFDs).^1^ Electrical stimulation of peripheral nerves is an effective third line treatment for OAB and PFD symptoms. One of the most common electrical stimulation therapy forms is sacral neuromodulation (SNM).^2^ SNM uses low amplitude current from a quadripolar electrode lead placed in a sacral foramen (usually S3 or S4) to modulate nerve activity with the goal of returning normal pelvic organ function.^3^ SNM systems have been used off label to stimulate the pudendal nerve when SNM has failed to relieve symptoms or in indications where pudendal nerve stimulation (PNS) may be preferable, such as pelvic pain.^4–7^ Branches of the S2-S4 sacral nerve roots join to form the pudendal nerve trunk (FIGURE 1), which splits distally into the perineal, inferior rectal, and dorsal genital nerves.^8^ The S2-S4 sacral roots also innervate the pelvic floor and lower limbs and contribute autonomic fibers to the pelvic nerve. Although they share many neural pathways, preclinical studies have suggested that SNM and PNS may have different mechanisms and act on the pelvic floor differently, but this remains unvalidated in humans.^9^

**Figure 1:**
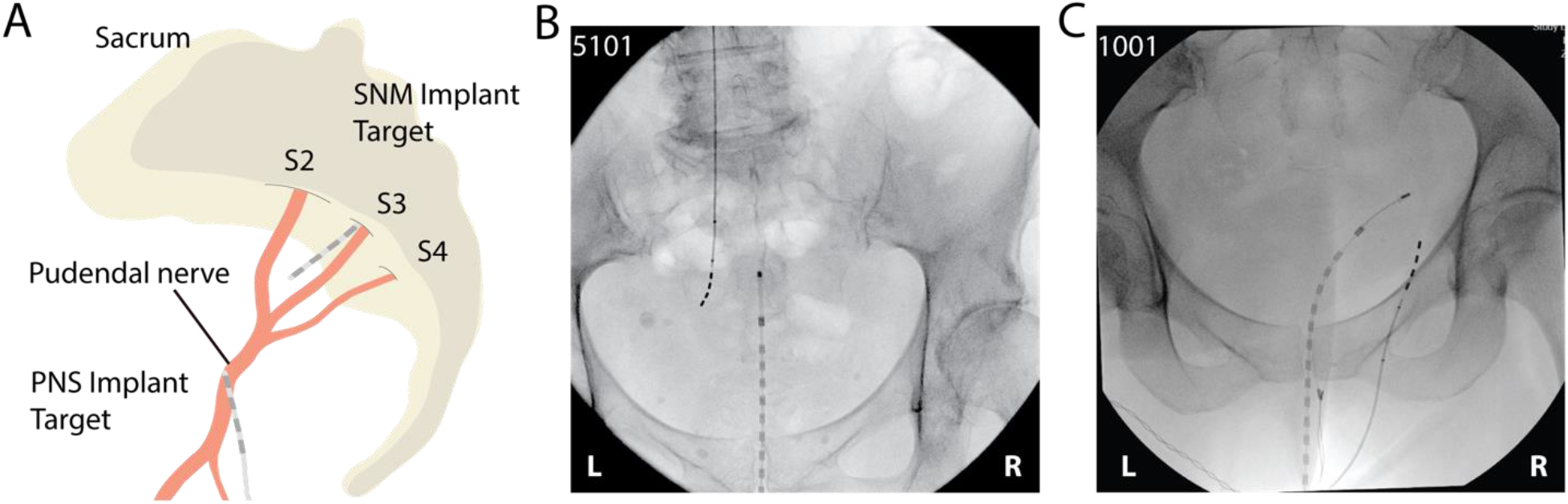
A: Illustration depicting the sacrum and sacral nerves S2-S4 and how they typically consolidate into the pudendal nerve. The most common nerve targets for SNM and PNS stimulation lead placement are depicted. B: Anteroposterior intra-operative fluoroscopy depicting an SNM lead placement (left side) and the intraurethral manometry catheter (center). C: Anteroposterior intra-operative fluoroscopy depicting a PNS lead placement (right side), the intraurethral manometry catheter (center), and EAS EMG needle placement (lower center). Each fluoroscopy image was obtained prior to the lead being tunneled and connected to a pulse generator.

Recently, our group used high-resolution manometry to characterize the intra-operative effect of PNS across the urethra.^10^ This work examined variations in urethra recruitment at different intensities of PNS and provides a framework with which to investigate sacral nerve stimulation (SNS). Several studies have investigated the intra-operative effect of SNS or PNS on the pelvic floor.^10–14^ No studies have directly compared pelvic floor responses between these two stimulation approaches. We collected intra-operative, SNS-evoked urethra and external anal sphincter (EAS) responses using high-resolution manometry and needle electromyography (EMG), respectively. We then compared SNS-evoked responses with previously collected PNS-driven responses data. This study is the first to investigate high-resolution urethral manometry during SNS and highlights how evoked responses of the pelvic floor may be used to compare recruitment characteristics of two similar neuromodulation modalities in humans.

## 2 Materials and Methods

This study was reviewed and approved by the Michigan Medicine Institutional Review Board and the Trinity Health Chelsea Hospital Institutional Review Board.

### 2.1 SNS participant recruitment

Nine female patients 18 years or older undergoing SNM implant operations at the Michigan Medicine main hospital (Ann Arbor, Michigan, USA) or the Trinity Health Chelsea Hospital (Chelsea, Michigan, USA) were assessed for eligibility and contacted for recruitment and consent. Potential participants with areflexive/atonic/neurogenic bladder or other conditions disrupting the micturition or storage circuits were excluded from eligibility.

### 2.2 SNS intra-operative set up

Each participant underwent the standard stage 1, or combined stage 1 and 2, SNM implant procedure (InterStim, Medtronic, Minneapolis, MN) in the prone position while under monitored anesthesia care. EMG needle electrodes (Ambu Neuroline, twisted pair subdermal, 74612– 250/1/20) were placed just lateral to the external anal sphincter (EAS) at the 3 and 9 o’clock positions. A multichannel manometry catheter (ManoScan MSC-3886, Medtronic) was inserted into the lower urinary tract such that at least two sensors were in the bladder and secured in place by taping the segment outside of the lower urinary tract (LUT) to the inside of the thigh.^10,15^ The manometry catheter is 8 Fr in diameter. Each of the 36 sensors on the catheter span 4 mm with a center-to-center distance of 7.5 mm.

### 2.3 Intra-operative SNS

Following standard procedures during lead placement, an external stimulator (InterStim Verify system, Medtronic) was used for intra-operative SNS. Once the SNM lead placement was finalized, we identified the visual motor threshold (VMT) for each lead electrode as the minimum current level required to elicit anal bellows or a toe twitch with 210 μs pulse width, 3 Hz monopolar stimulation. A low stimulation frequency was selected to observe the effect of each stimulation pulse on urethral pressure. Stimulation amplitude was increased from VMT to the highest current amplitude that would not lead to discomfort after the surgery, typically three or four times VMT. A set of SNS pulses at each multiple of VMT was considered a trial. If there was a response to stimulation in a segment of the urethra or EAS at any multiple of VMT for a given electrode, this was considered a successful recruitment attempt.

### 2.4 Urethral pressure data processing

Urethral pressure data from the manometry system was sampled at 100 Hz and processed using a custom MATLAB (version 2025a, MathWorks, Natick, MA) script. The span of the urethra was identified after reviewing the pressure profile within the LUT without stimulation. The proximal urethra channel was defined as the highest-pressure channel. The middle, middle/distal, and distal urethra channels were identified as the next three channels distal to the max pressure channel. This method is consistent with previous work.^10^ Each pressure signal was high-pass filtered at 1 Hz to remove breathing and movement artifacts. MATLAB’s “findpeaks” function was used to detect the pressure maximum produced by each stimulation pulse. We identified the change in pressure per stimulation pulse as the difference between the maximum pressure and the minimum pressure in the 250 ms preceding the maximum (ΔP; FIGURE 2). The urethral pressure response for each electrode and amplitude setting was calculated as the mean of ten consecutive ΔP values.

**Figure 2:**
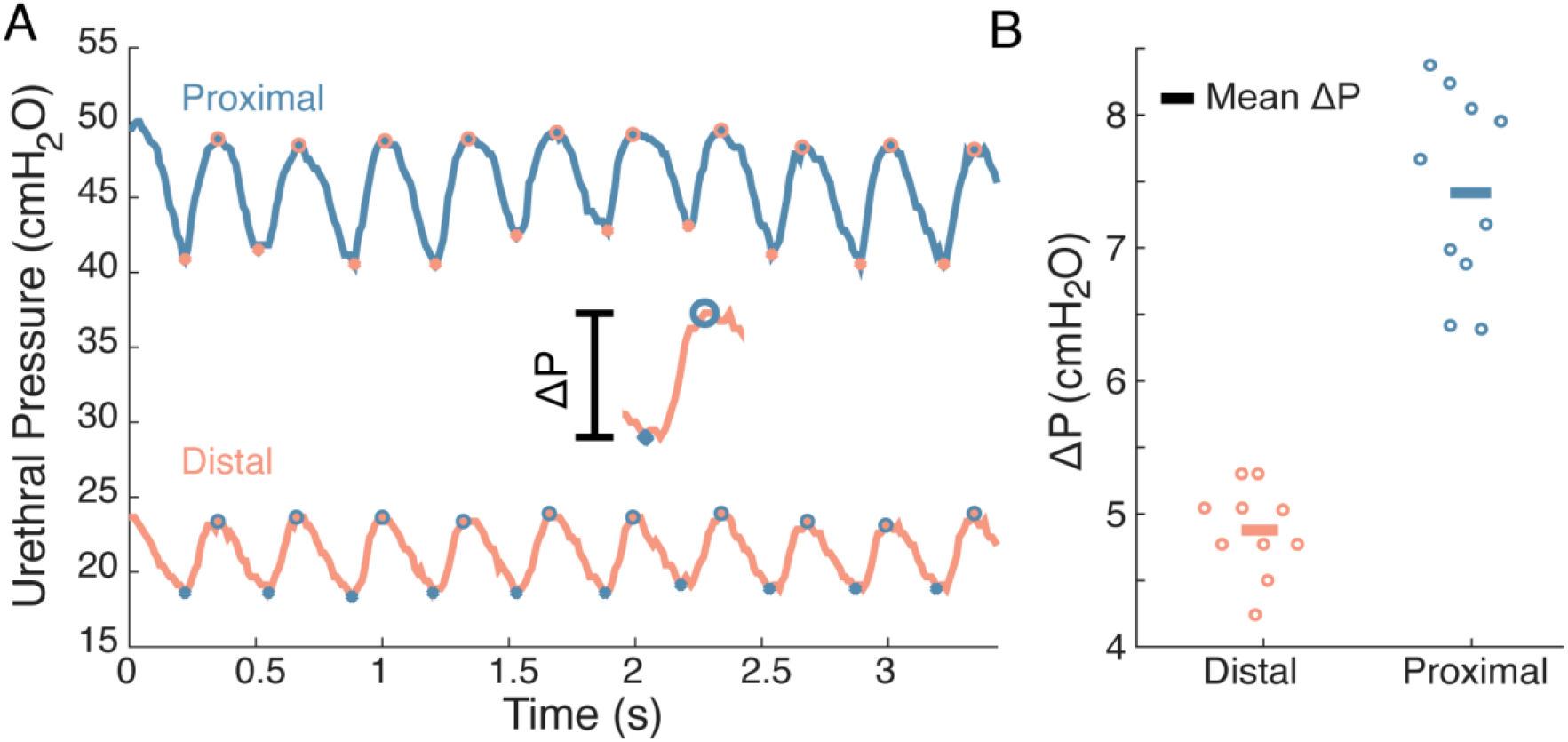
A: Example pressure signal in proximal and distal urethra locations during 3 Hz sacral nerve stimulation. For clarity, high-pass filtering was not used for this example. Local maxima are denoted by “o” and local minima as ‘.’. ΔP is the difference between each sequential local minima and subsequent maxima. B: ΔP values from the data in A. Mean ΔP for this example data is denoted by a horizontal line.

### 2.5 EAS EMG data processing

EAS EMG was collected at 10 kHz using a bioamplifier and recording module (ADInstruments Quad Bio-Amplifier and PowerLab, Colorado Springs, CO). EMG data was exported to a text file and imported into MATLAB (version 2025a, MathWorks, Natick, MA) for processing using a custom script. After time-aligning the EMG to the pressure data and segmentation into trials, the “findpeaks” function was used to locate ten consecutive stimulation artifacts associated with each trial. Stimulus triggered averaging was then employed to obtain the mean EMG response within the 2 to 100 ms interval post-stimulus in each trial. This mean response was rectified and integrated to obtain an area-under-the-curve (AUC) measure for each trial.

### 2.6 PNS data acquisition

PNS intra-operative data collection and data processing followed similar steps and is previously described by Chen et al. and Lagunas et al.^10,12^ For comparison to SNS data collected in this study, we only included responses to PNS that were elicited with 3 Hz and 210 μs pulse width monopolar stimulation. This led to the inclusion of intra-operative data from 11 patients receiving a PNS implant (TABLE S1).

### 2.7 Statistical analysis

Data from SNS and PNS experiments were analyzed in excel (version 16.106, Microsoft, Seattle, WA) and R (version 4.4.1, Vienna, Austria). The maximum mean ΔP due to SNS was compared across the urethra using a one-way analysis of variance (ANOVA). The distribution of electrodes producing the maximum mean ΔP, for SNS and PNS, was analyzed using a chi-square test for homogeneity. The urethra and EAS response to SNS across VMTs for each of the four electrodes was assessed by normalizing to each participant’s maximum response produced in each recording location. Relationships between VMTs and electrodes were tested using a two-way ANOVA. The maximum mean ΔP, current at maximum ΔP, minimum current to activate each urethra segment, and the mean ΔP per change in current for SNS and PNS were each compared using a two-way ANOVA. The mean current threshold for SNS activating any urethra segment or EAS was identified and compared to VMT using a t-test. The mean current threshold for activating each urethra segment and the EAS across participants and electrodes were identified and compared between recording locations, urethra segments and EAS, and between SNS and PNS using a two-way ANOVA. When relevant, data is reported as mean ± standard deviation. All statistical testing used a significance level of α = 0.05.

## 3 Results

### 3.1 Maximum SNS evoked urethral pressure change was higher in the distal urethra

In TABLE 1, we report the maximum mean ΔP due to SNS in the urethra for each participant. The mean ΔP in the middle/distal and distal urethra evoked by SNS was higher (4.7 and 3.6 cmH_2_O) than in the proximal and middle urethra (2.7 and 3.2 cmH_2_O), albeit not significantly (One way ANOVA, p = 0.6). The distribution of which electrode evoked the max response was significantly different than a uniform distribution (p = 0.006, TABLE 2), with E3 the most common electrode producing the maximum response. The electrodes E3 and E2, further from the end of the lead, typically evoked larger responses than distal electrodes (FIGURE 3).

**TABLE 1:**
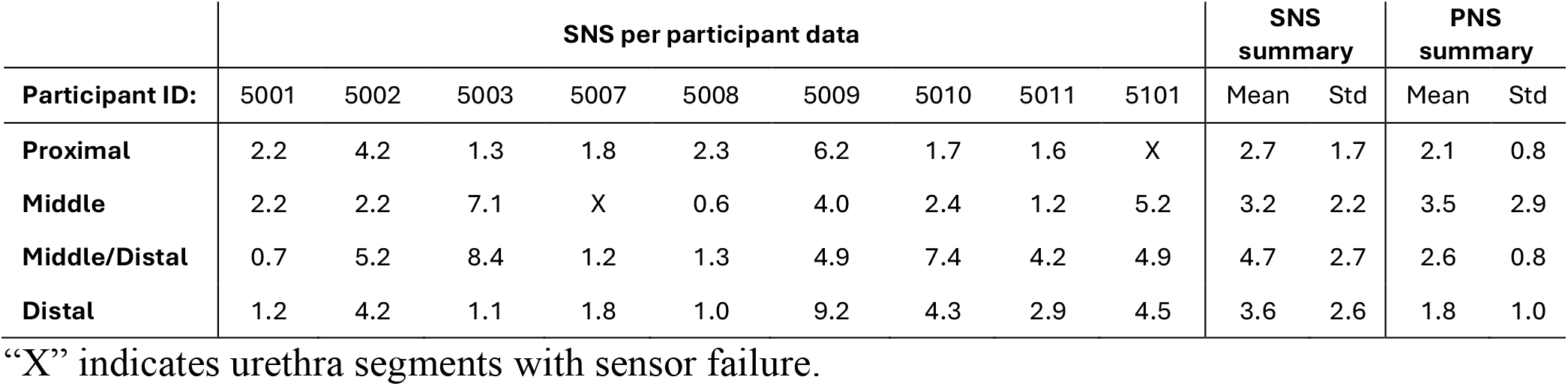
Maximum mean ΔP for each urethra segment in SNS (cmH_2_O)

**TABLE 2:** Distribution of electrodes driving the maximum mean ΔP.

|  | **E3** | **E2** | **E1** | **E0** | **Chi-square test for homogeneity (p-value)** |
|---|---|---|---|---|---|
| **SNS** | 15/34 | 12/34 | 4/34 | 3/34 | 0.006 |
| **PNS** | 0/37 | 9/37 | 10/37 | 18/37 | 0.0005 |

**Figure 3:**
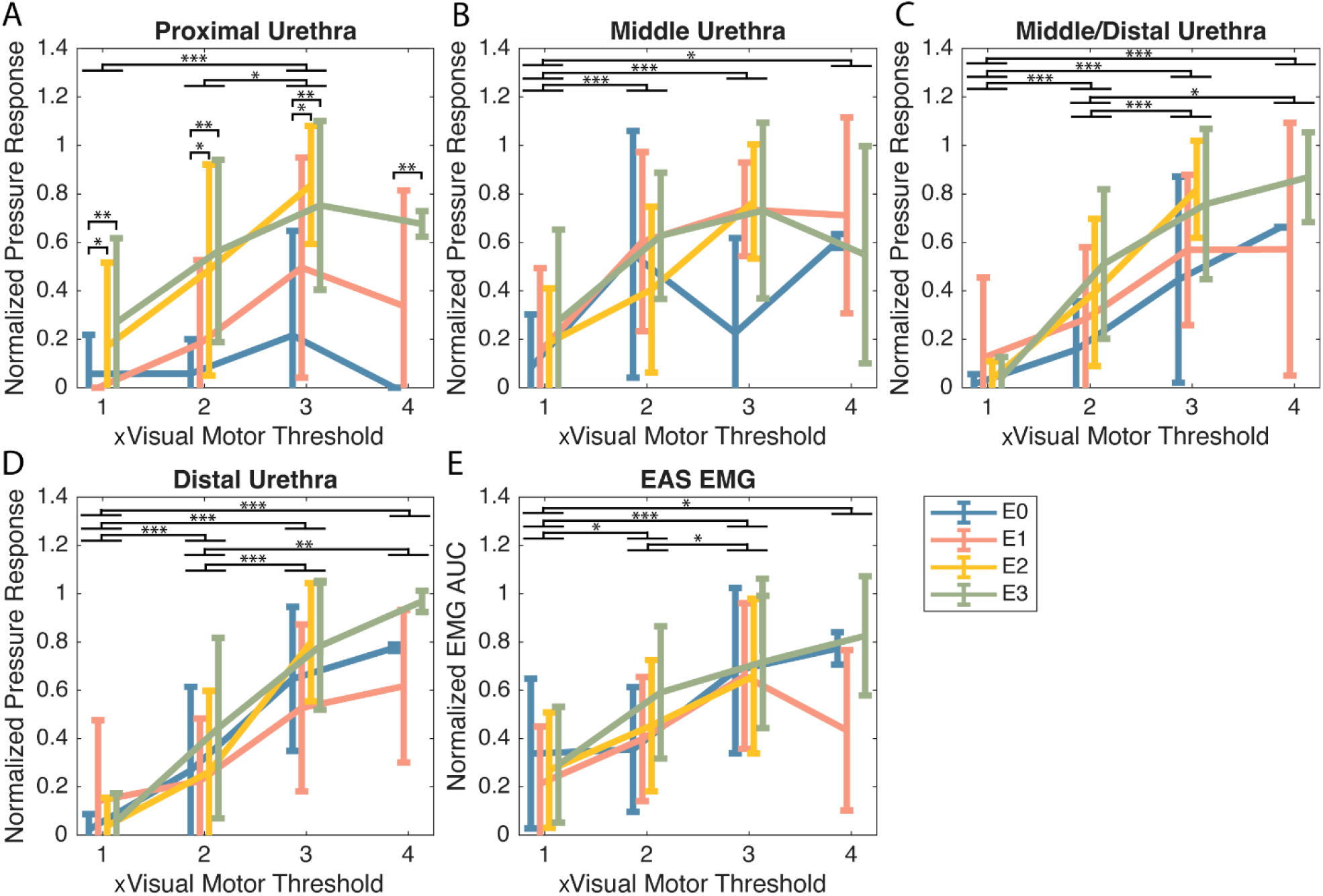
Mean normalized urethral and anal sphincter recruitment during intra-operative SNS across all SNS participants for each monopolar stimulation configuration (E0 – Blue, E1 – Salmon, E2 – Yellow, E3 – Green). Each participant’s response is normalized to the max response in that segment of the urethra (A-D) or to their max EAS response (E). Error bars are one standard deviation. *: p < 0.05, **: p < 0.005, ***: p< 0.0005 (two-way ANOVA followed by post-hoc Tukey test).

### 3.2 There was no significant difference between evoked max mean ΔP in pudendal and sacral nerve stimulation

TABLE 1 reports the maximum mean ΔP due to PNS, adapted from Chen et al. 2025^10^; per-participant measures are given in TABLE S1. PNS evoked higher pressures in the middle and middle/distal urethra (3.5 and 2.6 cmH_2_O) than in the proximal and distal urethra (2.1 and 1.8 cmH_2_O). During PNS the distribution of which electrode elicited the max response was also significantly different than a uniform distribution (p = 0.0005). E0 was the most common PNS electrode for producing the maximum response (TABLE 2). There were no significant differences in evoked maximum urethra mean ΔP between PNS and SNS (Two-way ANOVA, p = 0.09). There was a significant difference between the current amplitudes required to elicit maximum mean ΔP between SNS and PNS (3.0 ± 1.5 vs 3.8 ± 1.1 mA respectively, TABLE S2; Two-way ANOVA, p = 0.01). A post-hoc Tukey test revealed no significant differences between individual segments of the urethra but did confirm a significant difference between SNS and PNS (p = 0.01). There was a significant difference between SNS and PNS when comparing maximum mean ΔP per mA (1.5 ± 1.4 vs 0.8 ± 0.6 cmH_2_O per mA, respectively, TABLE S3; p = 0.004). Similarly, a post-hoc Tukey test revealed no significant differences between individual urethra segments (all p > 0.60) but showed a significant difference between stimulation modalities (p = 0.005). We repeated these analyses with the minimum current required to elicit a measurable change in pressure in each segment of the urethra and found no significant differences in change in pressure, current amplitudes, or cmH_2_O per mA in SNS versus PNS (Two-way ANOVA, p = 0.96, 0.32, and 0.59 respectively).

### 3.3 Recruitment order of pelvic muscles was similar between SNS and PNS

Across participants and SNS electrodes, the mean VMT was 1.7 ± 1.3 mA, the mean current threshold to activate any segment of the urethra was 2.5 ± 2.0 mA, and the mean current threshold to activate the EAS was 2.3 ± 1.2 mA. The mean current threshold to activate any urethra segment was significantly higher than VMT, though the mean current threshold of EAS activation was not (Welch two-sample t-test, p = 0.04 and p = 0.06 respectively). For previously collected PNS-driven data, the mean EAS threshold was 1.6 ± 1.1 mA. A two-way ANOVA showed no significant difference in current required to drive a urethra or EAS response between SNS and PNS (TABLE S4; p = 0.3) but did find a significant difference between recording locations (p = 0.04). A post-hoc Tukey test did not yield a significant difference between locations (minimum p = 0.08 across the EAS to urethra comparisons). SNS also had a higher percentage of successful recruitment attempts in each recording location, except for the proximal urethra (TABLE S4).

## 4 Discussion

This work represents the first study to directly compare urethral pressure and EAS EMG responses evoked by SNS and PNS in humans. There was no significant difference in max mean ΔP along the urethra in SNS or in PNS, nor between the two stimulation modalities. PNS required more current to reach the maximum ΔP. The recruitment order of SNS and PNS was similar, with no significant differences in thresholds between the two stimulation modalities.

This data shows that while PNS and SNS may start to recruit pelvic floor muscles at similar thresholds, SNS is more efficient in eliciting a larger response past threshold per mA of applied current.

SNS had a non-significant trend of a larger pressure response in the distal urethra than proximal urethra (TABLE 1). This finding is of interest as PNS yielded the opposite trend (TABLE 1). It has not previously been shown in humans that SNS and PNS may have differences in how they facilitate neural control of the urethra. A possible explanation is that SNS-driven responses are affected by the activation of efferent autonomic fibers forming the pelvic nerve. Previous work using atropine sulfate to inhibit central muscarinic cholinergic pathways in preclinical models has shown that efferent pelvic nerve activation promotes relaxation of the proximal urethra.^16^ Pelvic nerve activation in SNS, in combination with activation of somatic pudendal efferent fibers in both SNS and PNS, may account for the difference in urethral recruitment trends between SNS and PNS.

It is not unexpected that the electrode furthest from the end of the SNM lead evoked the maximum pressure most often (TABLE 2). Previous intra-operative response studies have shown that electrodes further from the end of the lead produced the lowest sensory and motor thresholds.^14,17^ Additionally, in large preclinical models E3 had the lowest motor threshold of the four electrodes.^18^

Current amplitudes required to reach maximum responses and the mean ΔP per mA were significantly different between SNS and PNS. The difference in required current to reach a maximum response could be due to the S3 nerve root containing motor units for pelvic floor muscles that the pudendal nerve does not,^19^ such as the levator ani, thereby increasing pressure greater than just EUS activation alone. The neuromodulation lead may also be closer, or more directly aligned, to the nerve in SNS than in PNS, leading current amplitudes and the ΔP per mA to be significantly different (TABLES S2 and S3, respectively). While our observation of no difference in minimum applied current to drive ΔP between SNS and PNS may challenge this idea, previous literature reporting electrode to nerve distances support this assessment. InterStim placement performed on cadavers documented the furthest electrode from nerve at a median distance of 3.5 mm;^20^ whereas, in a study with PNS recipients that utilized MRI and CT imaging, the furthest electrodes were between 5 mm and 20 mm away from the nerve.^12^ A limitation of this comparison is that these studies used different approaches to measure the distance, cadaver dissections and clinical imaging, affecting the reliability of the comparison. Ultimately this difference in required current to reach a similar max pressure response has clinical implications because often the first or second troubleshooting step is to increase the amplitude of stimulation.^21^ Our data suggests that this increase in stimulation may be more effective in SNS than PNS.

Recently, it has been suggested that the mechanisms of SNM involve sub-sensory activation of pelvic floor muscles, generating afferent input to the spinal cord.^3^ SNM programming is normally set just under the sensory threshold,^22^ which has been reported as up to 60 percent above VMT (1.23 vs. 0.94 mA in SNM responders^23^, 1.1 vs. 0.7 mA^24^, 0.87 vs. 0.73 mA^25^, sensory threshold vs VMT respectively). In this study, although VMT was approximately twice as high as in previous work, there was often no urethra activation at VMT (FIGURE 3). Despite this lack of activation at VMT, our mean EAS and urethra thresholds were 35 and 47 percent higher (0.6 and 0.8 mA higher), respectively, than VMT, which is within the range of previous SNM studies’ relative ratios. These results support that subsensory activation of the urethra and/or EAS pathways are involved in SNM mechanisms.

The primary limitation of this work was that data for SNS and PNS comparisons were collected from different participants. An ideal study design would involve collecting SNS and PNS data from the same individuals, however few individuals receive both implants. Another major limitation of this study was low recruitment; however, a sample size of nine for one stimulation modality and twenty total participants is comparable with contemporary intra-operative neural stimulation studies.^11,13,24^ Finally, using intervals of VMT as the step-size for current reduced the resolution at which we could observe responses to stimulation. This method was used to limit the duration of data collection and minimally impact normal surgical procedures. In the future, MRI-CT co-registration imaging studies could be performed to determine the relative distance of SNM lead electrodes to the sacral nerve target.

## 5 Conclusion

This study is the first to investigate differences in evoked pelvic floor responses between SNS and PNS. Our observations suggest that activation of pelvic nerve fibers during SNS may cause the urethra to respond differently than to PNS. SNS requires significantly lower amplitudes of current to produce maximum urethra pressures than PNS and has greater urethra recruitment per mA than PNS. This suggests that the SNS lead may be closer to the nerve target than in PNS and that using increases in stimulation amplitude to improve therapeutic response may be more effective in SNS than PNS. Future studies investigating direct stimulation effects should incorporate MRI-CT co-registration to determine SNM lead to nerve target-distances.

## Data Availability

All data produced in the present study are available upon reasonable request to the authors.

## Acknowledgements

The authors thank Yike Liu for assistance with subject recruitment. We also thank Sonia A. Bhaskaran and Amador C. Lagunas for assistance with data collection and Luis Ruiz for assistance in creation of Figure 1A. The authors would also like to thank Jessica A. Fagan for assistance proofreading the article at various stages.

