## Supplemental for "Sacral nerve stimulation is more efficient in facilitating pelvic floor responses than pudendal nerve stimulation"

**TABLE S1:** Maximum mean ΔP for each urethra segment in PNS ($cmH_2O$), adapted from [10]

| Participant ID: | 1001 | 1002 | 1007 | 1012 | 1016 | 1017 | 1019 | 1020 | 1021 | 1022 | 1024 | Mean | Std |
|---|---|---|---|---|---|---|---|---|---|---|---|---|---|
| Proximal | 2.4 | 2.4 | 3.2 | 2.3 | 1.0 | 2.3 | 1.4 | 2.8 | 1.8 | 18.3* | 1.2 | 2.1 | 0.8 |
| Middle | 1.8 | 1.7 | 4.4 | 1.9 | 0.9 | 1.7 | 3.9 | 10.6 | | 5.3 | 2.3 | 3.5 | 2.9 |
| Middle/Distal | 2.5 | 2.7 | 3.6 | | 1.1 | 2.8 | 3.0 | 3.4 | | 1.9 | 2.0 | 2.6 | 0.8 |
| Distal | | 1.4 | | | 0.7 | 3.7 | 2.3 | 0.9 | | 2.3 | 1.4 | 1.8 | 1.0 |

Blank spaces indicate where there was no urethra response. "*" indicates removal from statistical analysis due to being greater than three standard deviations above the mean.

**TABLE S2:** Current at maximum mean ΔP for each urethra segment in SNS and PNS (mA)

| Mean ± Std | Proximal | Middle | Middle/Distal | Distal | All urethra segments |
|---|---|---|---|---|---|
| SNS | 2.9 ± 1.5 | 2.4 ± 1.3 | 3.3 ± 1.6 | 3.2 ±1.6 | 3.0 ± 1.5 |
| PNS | 3.7 ± 1.1 | 3.8 ± 1.2 | 3.9 ± 1.3 | 3.7 ±0.9 | 3.8 ± 1.1 |

**TABLE S3:** Maximum mean ΔP per mA increment for each urethra segment in SNS and PNS ($cmH_2O$ per mA)

| Mean ± Std | Proximal | Middle | Middle/Distal | Distal | All urethra segments |
|---|---|---|---|---|---|
| SNS | 1.4 ± 1.3 | 1.7 ± 1.6 | 1.6 ± 1.4 | 1.4 ± 1.5 | 1.5 ± 1.4 |
| PNS | 0.6 ± 0.4 | 1.1 ± 0.9 | 0.7 ± 0.4 | 0.5 ±0.5 | 0.8 ± 0.6 |

**TABLE S4:** Mean current required for a response at each recording location (mA)

| | SNS | | PNS | |
|---|---|---|---|---|
| Recording location | Mean ± Std | Attempts with successful recruitment (%) | Mean ± Std | Attempts with successful recruitment (%) |
| EAS | 2.3 ± 1.2 | 94 | 1.6 ± 1.1 | 86 |
| Proximal Urethra | 2.2 ± 1.0 | 58 | 2.7 ± 1.5 | 61 |
| Middle Urethra | 2.6 ± 2.0 | 78 | 2.6 ± 1.6 | 61 |
| Middle/Distal Urethra | 2.8 ± 1.9 | 78 | 2.4 ± 1.4 | 57 |
| Distal Urethra | 2.7 ± 1.4 | 75 | 2.6 ± 1.3 | 50 |
